# Restorative Justice Approaches to Conflict Management in Healthcare Workplaces: A Systematic Review Protocol

**DOI:** 10.1101/2025.10.01.25337131

**Authors:** Ravi Shankar, Fiona Devi, Xu Qian

**Author notes:** **Corresponding Author:** Dr Ravi Shankar; Research and Innovation, Medical Affairs, Alexandra Hospital, Singapore Email correspondence.

## Abstract

**Background:** Healthcare workplaces experience significant interpersonal conflicts affecting staff wellbeing, patient safety, and organizational performance. Traditional punitive approaches to conflict management often fail to address underlying issues, potentially perpetuating cycles of dysfunction. Restorative justice, emphasizing healing, accountability, and relationship repair over punishment, offers promising alternatives for healthcare conflict resolution. Despite growing implementation, systematic evidence synthesis regarding effectiveness, implementation factors, and outcomes remains absent.

**Objectives:** This systematic review protocol aims to synthesize evidence on restorative justice approaches for managing workplace conflicts in healthcare settings, examining implementation processes, effectiveness, barriers, facilitators, and impacts on staff wellbeing, patient care, and organizational culture.

**Methods:** Following PRISMA-P guidelines, we will search ten databases (PubMed, MEDLINE, CINAHL, PsycINFO, Embase, Scopus, Web of Science, Business Source Premier, Cochrane Library, and ProQuest) from inception to December 2025. The SPIDER framework guides eligibility criteria focusing on healthcare workers involved in restorative justice interventions, their experiences and outcomes across diverse healthcare contexts. Covidence will facilitate study screening and selection. Quality assessment will employ the Mixed Methods Appraisal Tool (MMAT), with risk of bias evaluated using appropriate domain-specific tools. Narrative synthesis and thematic analysis will integrate quantitative and qualitative findings. GRADE-CERQual will assess confidence in qualitative evidence synthesis.

**Discussion:** This protocol anticipates generating comprehensive evidence regarding restorative justice implementation models, effectiveness indicators, contextual factors influencing success, stakeholder experiences, and comparative advantages over traditional approaches. Evidence generated will inform policy development, implementation guidelines, and training programs for healthcare organizations seeking transformative conflict resolution approaches that prioritize healing, learning, and relationship restoration over punitive measures.

## Introduction

Healthcare workplaces represent complex social environments where interpersonal conflicts arise from multiple sources including hierarchical tensions, professional boundary disputes, resource constraints, ethical disagreements, and communication breakdowns [1]. These conflicts profoundly impact healthcare delivery, with research demonstrating associations between workplace conflict and increased medical errors, reduced patient satisfaction, staff turnover, burnout, and organizational dysfunction [2]. Traditional approaches to healthcare conflict management, rooted in punitive human resources frameworks emphasizing investigation, blame attribution, and disciplinary action, often fail to address underlying systemic issues while potentially exacerbating interpersonal tensions and organizational culture problems [3]. Recent studies indicate that many healthcare workers experience workplace conflict on a regular basis, with some reporting it as a daily occurrence. [4].

The healthcare sector faces unprecedented challenges in maintaining positive workplace cultures amid increasing demands, resource constraints, and workforce shortages. These conflicts range from interprofessional tensions and bullying to discrimination, ethical disagreements, and leadership disputes. The COVID-19 pandemic intensified workplace stressors, with healthcare workers reporting increased conflicts related to resource allocation, safety protocols, vaccination policies, and moral distress [5]. Traditional conflict management approaches, typically involving formal grievance procedures, investigations, and disciplinary actions, consume substantial organizational resources while often leaving parties dissatisfied and relationships damaged [6]. Research indicates that formal grievance procedures can escalate conflicts, damage relationships permanently, and fail to address systemic issues contributing to workplace dysfunction [7].

Restorative justice represents a paradigm shift from punitive to healing-centered approaches, emphasizing accountability, understanding, and relationship repair rather than punishment and blame [8]. Originating in criminal justice contexts, restorative justice principles have been adapted for organizational settings, including healthcare, offering structured processes for addressing harm, facilitating dialogue, and rebuilding trust [9]. Core restorative justice principles include voluntary participation, facilitated dialogue between affected parties, focus on understanding harm and its impacts, collaborative problem-solving, and commitment to behavioral change and relationship repair [10]. Healthcare applications range from addressing interpersonal conflicts and bullying to managing patient complaints and adverse events. Evidence from pilot implementations suggests that restorative approaches can reduce repeat conflicts, improve workplace relationships, and enhance organizational culture [11].

Despite increasing adoption of restorative justice approaches in healthcare organizations internationally, systematic evidence regarding implementation processes, effectiveness, and outcomes remains fragmented. Existing literature consists primarily of case studies, pilot evaluations, and theoretical discussions lacking systematic synthesis [12]. Questions persist regarding optimal implementation models, training requirements, cultural prerequisites, outcome measurement, and comparative effectiveness versus traditional approaches. Healthcare leaders require evidence-based guidance for determining when restorative approaches are appropriate, how to implement them effectively, and what outcomes to expect. The absence of synthesized evidence hampers informed decision-making about adopting restorative justice approaches and limits understanding of factors contributing to successful implementation.

This systematic review addresses critical knowledge gaps by synthesizing diverse evidence sources examining restorative justice applications in healthcare workplace conflict management. The review adopts a broad perspective encompassing various restorative practices including facilitated conversations, peace circles, mediation with restorative elements, and formal conferencing. By examining implementation processes, stakeholder experiences, and outcomes across different healthcare contexts, this synthesis will provide comprehensive understanding of how restorative justice principles translate into healthcare workplace applications. The review’s mixed-methods approach recognizes that understanding complex organizational interventions requires integration of quantitative effectiveness data with qualitative insights about implementation processes and contextual factors.

### Objectives

This systematic review aims to synthesize evidence on restorative justice approaches for managing workplace conflicts in healthcare settings. The primary objective is to examine the effectiveness of restorative justice interventions compared to traditional conflict management approaches in healthcare workplaces. Secondary objectives include identifying and characterizing different restorative justice models implemented in healthcare settings and exploring factors that facilitate or hinder their successful implementation. The review will also synthesize stakeholder experiences to understand how healthcare workers, managers, and patients perceive and engage with restorative justice processes.

The review addresses several key research questions. The primary research question asks: What is the effectiveness of restorative justice approaches for managing workplace conflicts in healthcare settings compared to traditional disciplinary or grievance-based approaches? Secondary questions explore: What restorative justice models have been implemented in healthcare workplaces and what are their core components? What factors facilitate or hinder successful implementation of restorative approaches in healthcare organizations? How do different stakeholders experience restorative justice processes for workplace conflict resolution? What types of healthcare workplace conflicts are most amenable to restorative approaches? These questions align with growing recognition that transforming healthcare workplace cultures requires alternatives to traditional punitive approaches that often perpetuate dysfunction rather than promoting healing and learning [13].

## Methods

### Study Design and Protocol Registration

This mixed-methods systematic review protocol follows the Preferred Reporting Items for Systematic Reviews and Meta-Analyses Protocols (PRISMA-P) 2015 guidelines, ensuring comprehensive and transparent reporting of planned methods [14]. The protocol will be registered with the International Prospective Register of Systematic Reviews (PROSPERO) to promote transparency, reduce duplication, and minimize reporting bias. The review will employ the Joanna Briggs Institute (JBI) methodology for mixed-methods systematic reviews, enabling integration of quantitative, qualitative, and mixed-methods evidence [15]. Any protocol amendments during the review process will be documented with rationales and potential impacts on findings, maintaining transparency throughout the research process. The mixed-methods approach recognizes that understanding complex organizational interventions requires diverse evidence types capturing both effectiveness outcomes and implementation processes.

### Conceptual Framework

The SPIDER framework (Sample, Phenomenon of Interest, Design, Evaluation, Research type) structures this systematic review, providing a comprehensive approach for mixed-methods evidence synthesis [16]. This framework accommodates the methodological diversity expected in restorative justice research while maintaining systematic rigor. Unlike frameworks designed primarily for quantitative intervention studies, SPIDER’s flexibility enables capture of complex implementation processes, contextual factors, and diverse outcomes characterizing organizational interventions. The Sample component encompasses all healthcare workers involved in or affected by restorative justice interventions for workplace conflict, including clinical staff such as physicians, nurses, and allied health professionals, support staff, administrators, and managers across all healthcare settings. The Phenomenon of Interest captures restorative justice approaches to workplace conflict management, including various models such as conferences, circles, and mediation, implementation processes, and contextual factors influencing adoption and effectiveness. The Design component includes all empirical study designs providing evidence on restorative justice effectiveness or implementation, recognizing that different designs contribute complementary insights. The Evaluation component examines diverse outcomes including conflict resolution, participant satisfaction, workplace culture, staff wellbeing, and patient care indicators, acknowledging that restorative justice impacts multiple levels from individual to organizational. The Research type encompasses quantitative, qualitative, and mixed-methods studies, recognizing that understanding complex interventions requires diverse evidence types capturing different aspects of implementation and impact.

The SPIDER framework’s strength for this review lies in its accommodation of heterogeneous evidence while maintaining systematic rigor. The framework ensures comprehensive capture of implementation complexity characterizing organizational interventions while providing clear structure for evidence synthesis. Its emphasis on phenomenon of interest rather than specific interventions aligns with restorative justice as a philosophy manifesting through diverse practices rather than standardized protocols. The framework’s attention to evaluation recognizes that restorative justice outcomes extend beyond traditional effectiveness metrics to include relational, cultural, and transformative impacts requiring diverse assessment approaches.

### Eligibility Criteria

The population encompasses healthcare workers of all professional categories working in any healthcare setting who have participated in, facilitated, or been affected by restorative justice interventions for workplace conflict. This includes clinical staff comprising physicians, nurses, allied health professionals, and technicians as well as support staff, administrators, managers, and healthcare leaders. Patients and families are included when they participate in restorative processes addressing workplace conflicts that affect care delivery. Studies focusing exclusively on patient-provider disputes without workplace conflict components will be excluded, as the review specifically targets interprofessional and organizational conflicts.

The phenomenon of interest includes any restorative justice approach implemented to address workplace conflict in healthcare settings. This encompasses restorative conferences bringing affected parties together for facilitated dialogue, peace circles drawing on indigenous justice traditions, restorative mediation incorporating healing and relationship repair principles, and hybrid approaches combining restorative with traditional elements. Training programs preparing staff for restorative practices are included when they report implementation outcomes. Studies must address workplace conflicts rather than patient care disputes or criminal matters to meet inclusion criteria.

All healthcare settings are eligible including hospitals, primary care facilities, long-term care institutions, mental health services, and emergency departments across all geographical locations. Studies from all countries will be included without language restrictions, recognizing that restorative justice practices emerge from diverse cultural contexts globally. The temporal scope extends from database inception to December 2025, capturing the full evolution of restorative justice applications in healthcare.

Study designs comprise all empirical methodologies providing primary data about restorative justice implementation or effectiveness. Quantitative designs include randomized controlled trials, quasi-experimental studies, cohort studies, and pre-post evaluations. Qualitative designs encompass phenomenology, grounded theory, ethnography, case studies, and action research. Mixed-methods studies will be included when components meet inclusion criteria. Theoretical papers without empirical data, conference abstracts without full text, and opinion pieces will be excluded.

**Table 1:** Inclusion and Exclusion Criteria.

| Criteria | Inclusion | Exclusion |
| --- | --- | --- |
| Population | Healthcare workers (clinical, support, administrative) involved in restorative justice processes; Patients/families when part of workplace conflict resolution | Studies without healthcare worker involvement; Exclusively patient-provider disputes |
| Phenomenon | Restorative justice approaches for workplace conflict (conferences, circles, mediation, facilitated dialogue); Training programs | Traditional disciplinary approaches only; Criminal justice applications |
| Context | All healthcare settings (hospitals, primary care, long-term care, mental health, | Non-healthcare settings |
|  | emergency services) |  |
| Study Design | All empirical studies with primary data (quantitative, qualitative, mixed-methods) | Theoretical papers; Opinion pieces; Abstracts only |
| Time Period | Database inception to December 2025 | None |

### Information Sources and Search Strategy

Comprehensive database searching will encompass ten major databases ensuring interdisciplinary coverage of health, social science, and management literature. Health and medical databases include PubMed/MEDLINE for biomedical literature, CINAHL for nursing and allied health perspectives, Embase for international coverage including European sources, the Cochrane Library for systematic reviews and controlled trials, and Web of Science for citation tracking and interdisciplinary content. Social science and management databases comprise PsycINFO for psychological and behavioral aspects, Scopus for broad interdisciplinary coverage, Business Source Premier for organizational and management perspectives, and ProQuest Central for comprehensive multidisciplinary content. Grey literature sources include ProQuest Dissertations and Theses Global, OpenGrey for European grey literature, Google Scholar with the first two hundred results reviewed for relevance, organizational websites of healthcare quality and safety organizations, and conference proceedings from restorative justice and healthcare management conferences.

The search strategy employs a four-concept approach developed through iterative refinement with a health sciences librarian experienced in systematic review searching. The strategy combines terms for restorative justice including “restorative justice,” “restorative practice,” “restorative approach,” “peace circle,” “healing circle,” and “facilitated dialogue” with healthcare setting terms such as “healthcare,” “hospital,” “clinic,” “medical,” “nursing,” and “health professional.” Conflict-related terms include “conflict,” “dispute,” “grievance,” “bullying,” “harassment,” “discrimination,” and “workplace violence.” Workplace context terms encompass “workplace,” “organizational,” “staff,” “employee,” “interprofessional,” and “team.” The strategy will be adapted for each database while maintaining conceptual consistency, with search strings customized for database-specific syntax and indexing terms. Supplementary search strategies include hand-searching reference lists of included studies and relevant systematic reviews, forward citation searching of key studies using Google Scholar and Web of Science, contacting experts in restorative justice and healthcare management for unpublished studies, and searching organizational websites implementing restorative justice programs.

### Study Selection

Covidence systematic review software will manage the entire selection process, providing systematic tracking and documentation of all screening decisions [17]. Two independent reviewers will conduct title and abstract screening followed by full-text assessment using standardized screening forms developed specifically for this review. Before commencing screening, reviewers will independently screen one hundred randomly selected citations in a calibration exercise, discussing discrepancies to ensure consistent application of eligibility criteria. Inter-rater reliability will be assessed using Cohen’s kappa statistics, with values of 0.75 or greater indicating acceptable agreement [18]. If initial agreement falls below this threshold, additional training and calibration will occur before proceeding with full screening.

Title and abstract screening will involve both reviewers independently assessing all identified citations against inclusion criteria, with studies marked as include, exclude, or maybe for further assessment. Full-text assessment will involve detailed evaluation against all eligibility criteria, with specific reasons for exclusion documented for transparency and reproducibility. Disagreements during screening will be resolved through discussion between the two reviewers, with a third reviewer consulted when consensus cannot be reached through discussion. Authors of studies will be contacted when additional information is needed to determine eligibility, with a maximum of three contact attempts over a four-week period before excluding studies due to insufficient information. A PRISMA flow diagram will document the complete selection process, including the number of citations identified from each source, citations excluded at each screening stage, reasons for exclusion at full-text screening, and final numbers of included studies, ensuring transparency and reproducibility of the selection process.

### Data Extraction

A standardized data extraction form developed specifically for this review will capture comprehensive information about study characteristics, methodological approaches, participant demographics, intervention details, and findings relevant to restorative justice in healthcare settings. The extraction form will be piloted on five randomly selected included studies and refined based on this pilot before full extraction begins. Two reviewers will independently extract data from each included study, with discrepancies resolved through discussion and verification against original sources. Where information is unclear or missing, study authors will be contacted for clarification with up to three contact attempts over four weeks.

Study characteristics to be extracted include authors, publication year, country and geographical setting, study aims and research questions, theoretical or conceptual frameworks employed, funding sources, and any declared conflicts of interest. Methodological information encompasses study design and methodology, sampling strategy and achieved sample size, data collection methods and timing, analysis approach including software used, strategies used to enhance trustworthiness or rigor, and ethical approval details. Participant characteristics will be documented including number and types of participants, professional roles and demographics such as age, gender, experience level, nature and severity of conflicts addressed, organizational context including size, type, and culture, and any relevant contextual factors influencing implementation or outcomes.

Detailed intervention information will capture the type of restorative justice approach used, core components and process elements, theoretical basis and adaptation from original models, facilitator training and qualifications, implementation timeline and intensity, comparison intervention characteristics if applicable, and any co-interventions or organizational supports provided. Outcomes and findings extraction will include primary and secondary outcomes measured, measurement tools and assessment timing, quantitative results including effect sizes and confidence intervals where available, qualitative themes with supporting participant quotations, implementation factors including barriers and facilitators, contextual influences on effectiveness, and unintended consequences or adverse events. Special attention will be given to extracting information about implementation processes, organizational readiness, culture change indicators, and sustainability factors that influence long-term adoption of restorative approaches.

### Quality Assessment

The Mixed Methods Appraisal Tool (MMAT) version 2018 will assess methodological quality across all study designs, providing a comprehensive framework for evaluating diverse evidence types [19]. Two reviewers will independently assess each study’s quality, with disagreements resolved through discussion and third reviewer consultation when necessary. The MMAT examines design-specific quality criteria while enabling comparison across different methodological approaches, making it ideal for mixed-methods systematic reviews.

For quantitative randomized controlled trials, assessment will examine randomization appropriateness including sequence generation and allocation concealment, baseline comparability between groups and management of confounding variables, completeness of outcome data and handling of attrition, blinding appropriateness where feasible given intervention nature, and adherence to intended intervention protocols. Non-randomized quantitative studies will be evaluated for participant representativeness and selection bias risk, measurement appropriateness and validation of outcome assessments, confounding control through design or analysis, intervention fidelity and contamination between groups, and completeness of outcome reporting including missing data handling.

Qualitative studies will be assessed for qualitative approach appropriateness for addressing research questions, adequacy of data collection methods for capturing phenomenon of interest, findings clearly derived from and grounded in data, interpretation coherence and sufficient data to support conclusions, and researcher reflexivity addressing potential influence on data generation and interpretation. Mixed-methods studies receive additional assessment examining rationale clarity for using mixed-methods design, effective integration of quantitative and qualitative components, adequate addressing of divergences between method findings, adherence to quality criteria for each component, and added value of integration beyond separate components.

Studies will be categorized as high quality when meeting eighty to one hundred percent of applicable criteria, moderate quality when meeting sixty to seventy-nine percent of criteria, or low quality when meeting fewer than sixty percent of criteria. Quality ratings will inform sensitivity analyses examining whether findings differ when lower quality studies are excluded, rather than serving as basis for study exclusion. This approach recognizes that even lower quality studies may provide valuable insights about implementation processes or contextual factors while ensuring that synthesis conclusions appropriately weight higher quality evidence.

### Risk of Bias Assessment

Risk of bias evaluation will employ design-specific tools providing detailed assessment of potential biases affecting study validity. For randomized controlled trials, the Cochrane Risk of Bias Tool 2.0 will examine bias arising from the randomization process including sequence generation and allocation concealment adequacy, deviations from intended interventions considering both assignment and adherence, missing outcome data including differential attrition between groups, measurement of outcomes including blinding and assessment standardization, and selective reporting comparing protocols with published results [20]. Each domain receives a judgment of low risk, some concerns, or high risk, with overall risk determined by the highest risk domain.

Non-randomized quantitative studies will be assessed using the ROBINS-I tool examining bias due to confounding including important confounders and their measurement, selection of participants including start of follow-up and selection methods, classification of interventions considering information sources and differential misclassification, deviations from intended interventions including co-interventions and contamination, missing data including differential missingness and inappropriate methods, measurement of outcomes including blinding and differential measurement error, and selection of reported results including multiple analyses and selective reporting [21]. Judgments range from low risk through moderate and serious to critical risk of bias, with overall assessment determined by the most serious risk level identified.

Qualitative studies will undergo critical assessment examining researcher reflexivity and positioning including background, relationship with participants, and potential influence on data generation; sampling strategy appropriateness for capturing diverse perspectives and achieving data saturation; ethical considerations including informed consent, confidentiality, and potential harm; rigor in analysis including systematic coding, member checking, and triangulation; and credibility of findings including thick description, negative case analysis, and peer debriefing. While qualitative research does not aim for the same type of objectivity as quantitative studies, these criteria ensure trustworthiness and credibility of findings contributing to the synthesis.

### Data Synthesis

Data synthesis will employ complementary approaches appropriate for integrating diverse evidence types while maintaining methodological rigor. The synthesis strategy recognizes that restorative justice represents a complex intervention requiring multiple synthesis methods to capture different dimensions of implementation and impact. Integration will occur at multiple levels from individual study findings through thematic patterns to overarching theoretical insights about how restorative justice functions in healthcare contexts.

Where studies examining similar interventions report comparable outcomes using consistent measures, meta-analysis will be conducted using random-effects models recognizing expected heterogeneity across healthcare contexts and populations. Statistical heterogeneity will be assessed using I-squared statistics, with values exceeding seventy-five percent indicating substantial heterogeneity requiring exploration through subgroup analyses or narrative synthesis rather than statistical pooling. Planned analyses include pooled effect sizes for common outcomes such as conflict resolution rates, participant satisfaction scores, and workplace culture indicators; subgroup analyses by healthcare setting, conflict type, intervention model, and implementation intensity; sensitivity analyses excluding lower quality studies to examine robustness of findings; and meta-regression exploring sources of heterogeneity including organizational size, implementation support, and facilitator training. Forest plots will display individual study effects and pooled estimates with confidence intervals, enabling visual assessment of consistency across studies.

Where meta-analysis proves inappropriate due to intervention heterogeneity, outcome diversity, or methodological differences, narrative synthesis will summarize and explain findings organized by outcome domain and intervention type. The narrative synthesis will follow Popay and colleagues’ guidance including developing a preliminary synthesis of findings, exploring relationships within and between studies, assessing synthesis robustness through sensitivity analyses, and developing a narrative explaining patterns and exceptions [22]. Tabulation will organize studies by key characteristics enabling pattern identification across contexts and populations. Vote counting based on direction of effect rather than statistical significance will provide an overall assessment of whether evidence suggests positive, negative, or null effects while avoiding inappropriate emphasis on individual study significance.

Qualitative synthesis will employ thematic synthesis following Thomas and Harden’s three-stage approach enabling systematic integration while preserving interpretive richness [23]. Line-by-line coding of findings sections will identify concepts and insights across studies, with initial coding remaining close to participants’ own words and authors’ interpretations. Descriptive themes will organize codes into patterns capturing common experiences, processes, and outcomes across studies while maintaining attention to divergent findings and contextual variations. Analytical themes will generate new interpretive insights extending beyond individual study findings to develop theoretical understanding of how restorative justice operates in healthcare contexts, what mechanisms produce change, and what conditions support effectiveness. NVivo qualitative data analysis software will facilitate systematic coding and theme development while maintaining an audit trail of analytical decisions.

Mixed-methods integration will employ the Pillar Integration Process enabling systematic combination of quantitative and qualitative syntheses [24]. The process involves listing findings from separate quantitative and qualitative syntheses organized by research question, matching findings to identify areas of convergence, complementarity, and divergence between evidence types, categorizing matched findings into pillars representing key insights supported by multiple evidence types, and assembling pillars into a coherent narrative addressing research questions while acknowledging evidence limitations. This approach ensures that different evidence types contribute their unique strengths while identifying where findings reinforce or challenge each other, ultimately producing a richer understanding than either synthesis alone could provide.

### Confidence Assessment

The Confidence in Evidence from Reviews of Qualitative research (CERQual) approach will assess confidence in synthesized qualitative findings based on four components [25]. Methodological limitations will be assessed by examining the extent to which studies contributing to each finding have methodological problems potentially affecting credibility based on MMAT assessments and qualitative criteria. Coherence evaluation determines whether findings are well-grounded in data from contributing studies and provide convincing explanations for patterns observed across contexts and populations. Data adequacy assessment examines whether studies provide sufficiently rich data to develop comprehensive understanding and whether the body of evidence contains enough studies to confidently support conclusions. Relevance appraisal evaluates the extent to which evidence from contributing studies applies to the review question’s context, population, and phenomenon of interest, considering whether findings transfer across different healthcare settings and conflict types.

Each synthesized finding will receive an overall CERQual confidence rating of high indicating minor concerns across all components suggesting finding is highly likely to be reasonable representation of phenomenon; moderate indicating moderate concerns in one or more components suggesting finding is likely reasonable representation but possibility of variation; low indicating substantial concerns in one or more components suggesting finding may be reasonable representation but substantial uncertainty exists; or very low indicating serious concerns across multiple components suggesting limited confidence in finding accuracy. Two reviewers will independently conduct CERQual assessments with disagreements resolved through discussion to reach consensus. Summary of Qualitative Findings tables will present each finding with its CERQual assessment and detailed explanation supporting the judgment, enabling users to understand evidence strength and make informed decisions about applicability to their contexts.

## Discussion

This systematic review protocol addresses critical gaps in understanding restorative justice applications for healthcare workplace conflict management at a time when healthcare organizations urgently seek alternatives to traditional punitive approaches that often perpetuate rather than resolve workplace dysfunction. The comprehensive methodology ensures rigorous synthesis while maintaining sensitivity to the complex, context-dependent nature of restorative interventions operating within diverse healthcare environments. By adopting mixed-methods approaches integrating quantitative effectiveness evidence with qualitative implementation insights, the review captures both whether restorative justice works and how it operates in practice, providing essential guidance for healthcare organizations considering adoption.

The protocol’s strength lies in its inclusive scope encompassing diverse restorative practices, healthcare settings, and conflict types, recognizing that restorative justice represents a philosophy and set of principles rather than a single standardized intervention. This breadth acknowledges that successful implementation requires adaptation to local contexts, organizational cultures, and specific conflict patterns rather than rigid adherence to prescribed protocols. The SPIDER framework provides appropriate structure for capturing this complexity while maintaining systematic rigor necessary for credible evidence synthesis. Unlike frameworks designed primarily for clinical interventions with standardized protocols and clearly defined outcomes, SPIDER accommodates the multifaceted nature of organizational interventions affecting multiple stakeholders through various mechanisms operating at individual, interpersonal, and system levels.

Healthcare organizations increasingly recognize that traditional punitive approaches to conflict management carry substantial hidden costs beyond direct resource consumption. Research indicates that formal grievance procedures often escalate conflicts, permanently damage working relationships essential for effective care delivery, fail to address systemic issues contributing to recurring conflicts, and create adversarial cultures undermining psychological safety necessary for quality improvement and patient safety [26]. The estimated annual cost of workplace conflict in healthcare exceeds twelve billion dollars in the United States alone when considering turnover, absenteeism, reduced productivity, and patient care impacts [27]. Restorative approaches offer potential for transformative culture change, shifting organizational focus from blame and punishment toward learning, healing, and relationship repair aligned with healthcare’s caring mission. However, implementation requires significant organizational commitment including leadership support, resource investment, culture change efforts, and sustained attention over time. This review will clarify what organizations can realistically expect from restorative justice implementation, helping leaders make informed decisions about adoption and resource allocation.

The review faces several anticipated challenges requiring proactive mitigation strategies. Terminology inconsistency represents a significant challenge as restorative practices may be described using various terms including “just culture,” “healing circles,” “accountability conferences,” or organization-specific names without using “restorative justice” language. The comprehensive search strategy addresses this through inclusive terminology, supplementary search methods, and expert consultation to identify relevant studies regardless of labeling. Intervention heterogeneity may limit possibilities for statistical meta-analysis given variation in restorative models, implementation approaches, and outcome measurement. The planned narrative and thematic syntheses ensure meaningful integration regardless of statistical pooling feasibility, with attention to identifying common principles and mechanisms across diverse practices. Publication bias toward positive results may overestimate effectiveness if unsuccessful implementations remain unpublished. Grey literature inclusion, quality assessment weighting, and sensitivity analyses help mitigate this risk while acknowledging that some implementation failures may remain undocumented.

The complex relationship between context and intervention effectiveness presents analytical challenges given that restorative justice success likely depends on organizational culture, leadership support, implementation quality, and conflict characteristics. The synthesis will explore these relationships through subgroup analyses, qualitative theme development, and mixed-methods integration while acknowledging that some contextual factors may remain poorly understood. Outcome heterogeneity reflects the multiple levels at which restorative justice operates, from individual wellbeing through team relationships to organizational culture, with different studies prioritizing different outcomes. The synthesis will organize outcomes by level and domain while examining whether effects at one level translate to impacts at other levels.

Findings will have immediate practical applications for healthcare organizations at various stages of considering or implementing restorative approaches. For organizations contemplating adoption, the synthesis will provide evidence-based guidance regarding readiness assessment including cultural prerequisites and leadership requirements, resource planning including training needs and facilitator development, implementation timelines with realistic expectations for observing changes, and risk assessment including potential challenges and mitigation strategies. Organizations currently implementing restorative approaches will benefit from evidence about optimization strategies for enhancing effectiveness, common implementation challenges and successful solutions, outcome metrics appropriate for tracking progress, and sustainability factors supporting long-term adoption. The synthesis will enable organizations to learn from others’ experiences rather than repeating common mistakes or reinventing successful strategies.

Healthcare systems internationally are reconsidering approaches to workplace culture and staff wellbeing, particularly following the COVID-19 pandemic’s revelation of workforce vulnerability and the critical importance of supportive work environments. This review will inform policy development at multiple levels from institutional through regional to national healthcare systems. Regulatory frameworks may need updating to recognize restorative approaches as legitimate alternatives to traditional disciplinary processes, establish standards for facilitator training and process quality, protect participants from retaliation while maintaining accountability, and integrate restorative options with existing professional regulation systems. Funding decisions require evidence about cost-effectiveness compared to traditional approaches, return on investment timelines, and resource requirements for successful implementation. The review will provide business cases supporting or tempering enthusiasm for restorative justice investment based on available evidence rather than anecdotal reports or theoretical promises.

Quality and safety implications extend beyond direct conflict resolution to broader impacts on care delivery. Psychological safety, recognized as essential for error reporting, quality improvement participation, and interprofessional collaboration, may be enhanced through restorative approaches emphasizing learning over blame [28]. The review will examine whether restorative justice implementation correlates with patient safety indicators, care quality metrics, and patient experience scores, providing evidence for or against claimed connections between workplace culture and care outcomes. These findings have implications for accreditation standards, quality improvement frameworks, and patient safety initiatives increasingly recognizing culture’s role in care delivery.

The review will identify critical knowledge gaps directing future research priorities in this emerging field. Effectiveness research needs include robust controlled trials comparing restorative and traditional approaches, long-term follow-up studies examining sustainability and culture change, economic evaluations including cost-effectiveness and return on investment analyses, and patient outcome studies linking workplace interventions to care quality. Implementation research priorities encompass optimal strategies for introducing restorative approaches in resistant cultures, adaptation requirements for different healthcare contexts and populations, factors supporting sustainability beyond initial enthusiasm, and approaches for scaling successful pilots to system-wide implementation. Mechanism research should explore how restorative processes produce observed changes, which intervention components are essential versus adaptable, what moderates effectiveness across contexts and conflicts, and how cultural factors influence acceptability and impact.

The synthesis will advance theoretical understanding of organizational conflict resolution in healthcare contexts, potentially extending existing frameworks from organizational psychology, implementation science, and healthcare management. Restorative justice challenges traditional organizational justice theories emphasizing procedural and distributive fairness by introducing relational and transformative dimensions previously undertheorized in healthcare management literature. The review may generate new theoretical insights about healing-centered organizational practices, contributing to emerging literature on trauma-informed organizational development gaining traction in healthcare and social services. These theoretical contributions extend beyond healthcare to inform understanding of restorative approaches in other high-stakes professional contexts characterized by hierarchical relationships, emotional intensity, and public accountability.

Methodological contributions include demonstrating rigorous approaches to synthesizing complex intervention evidence in organizational contexts. The integration of quantitative effectiveness data with qualitative implementation insights through systematic methods provides a model for reviewing organizational interventions where context profoundly influences outcomes. The careful attention to context-mechanism-outcome relationships through mixed-methods integration offers templates for future systematic reviews examining complex healthcare interventions operating at multiple levels. The protocol’s transparency about anticipated challenges and mitigation strategies contributes to methodological literature on conducting systematic reviews of emerging interventions with heterogeneous evidence bases.

Several limitations warrant acknowledgment to appropriately frame expectations and interpretation. The focus on peer-reviewed literature and formal grey literature sources may miss innovative practices implemented but not yet formally evaluated, potentially underestimating implementation prevalence. The broad scope, while comprehensive, may limit depth of analysis for specific applications such as particular conflict types or professional groups, suggesting need for subsequent focused reviews examining specific applications in greater detail. The review’s focus on workplace conflicts may miss insights from restorative justice applications in other healthcare contexts such as patient harm or medical error that could inform workplace applications. The heterogeneity of restorative justice interventions may complicate synthesis and limit possibilities for statistical meta-analysis, though planned narrative and thematic syntheses will ensure meaningful integration regardless of quantitative pooling feasibility. These limitations will be explicitly acknowledged in the review with implications for interpretation discussed.

Ethical considerations permeate the protocol despite involving synthesis of published research rather than primary data collection. Maintaining dignity and respect for participants in original studies requires careful contextualized use of quotations avoiding misrepresentation or stigmatization of individuals or organizations attempting innovative approaches. Equity considerations demand attention to whether evidence represents diverse populations and settings or reflects limited perspectives of well-resourced organizations in high-income countries. The review will explicitly acknowledge when evidence gaps exist for particular populations, settings, or conflict types, avoiding inappropriate generalization from limited evidence bases. Team reflexivity regarding members’ own experiences with workplace conflict and restorative justice will be maintained through regular debriefing sessions, ensuring that synthesis remains grounded in data rather than personal beliefs about optimal conflict management approaches.

Future implications extend beyond immediate evidence synthesis to shape the field’s development trajectory. Research priorities identified through the review will inform funding decisions and study design for next-generation restorative justice research in healthcare. Methodological insights about studying complex organizational interventions will improve future research quality, potentially leading to more rigorous and informative studies. Implementation guidance will accelerate adoption in organizations ready for change while preventing premature implementation in unprepared settings where failure might discredit restorative approaches more broadly. The review establishes foundation for evidence-based practice development in an emerging field currently characterized by enthusiasm exceeding evidence. By providing balanced assessment of what is known, unknown, and unknowable given current evidence, the synthesis enables informed decision-making while stimulating necessary research to address critical gaps.

The protocol’s comprehensive approach ensures that resulting synthesis will provide valuable guidance for multiple stakeholder groups navigating decisions about restorative justice adoption. Healthcare leaders will gain evidence-based frameworks for assessing organizational readiness, planning implementation, and evaluating outcomes. Practitioners involved in conflict resolution will understand which approaches show promise for different situations and what training and support they need for effective practice. Researchers will identify priority questions requiring investigation and appropriate methods for studying complex organizational interventions. Policymakers will have evidence to inform decisions about regulatory frameworks, funding priorities, and quality standards. Ultimately, healthcare workers experiencing workplace conflict may benefit from more effective, healing-centered approaches that restore relationships and improve workplace culture rather than perpetuating cycles of blame and dysfunction.

This systematic review protocol establishes rigorous foundation for synthesizing diverse evidence about restorative justice applications in healthcare workplace conflict management. The comprehensive methodology ensures systematic identification, evaluation, and integration of multiple evidence types while maintaining sensitivity to contextual factors profoundly influencing implementation and effectiveness. The mixed-methods approach recognizes that understanding complex organizational interventions requires integration of effectiveness data with implementation insights, stakeholder experiences, and contextual influences. By examining not just whether restorative justice works but how, when, for whom, and under what conditions, the review will generate nuanced understanding essential for successful translation into practice. The synthesis will contribute to transforming healthcare workplaces from environments often characterized by dysfunction, blame, and punishment into spaces supporting healing, learning, and collaborative problem-solving aligned with healthcare’s fundamental caring mission.

## Data Availability

All data produced in the present study are available upon reasonable request to the authors

